# Higher body mass index is a causal risk factor for skin infections: a Mendelian randomisation study using UK Biobank and FinnGen

**DOI:** 10.1101/2025.05.27.25328427

**Authors:** Rhian Hopkins, Ethan de Villiers, Michael N Weedon, Beverley M Shields, John M Dennis, Andrew P McGovern, Harry D Green

## Abstract

**Background:** Infections remain a leading cause of mortality and morbidity globally. Obesity is associated with increased infection risk in observational studies, however is it unclear if this association is causal. We used Mendelian randomisation to evaluate the potential causal role of higher BMI on common bacterial, viral, and fungal infections.

**Methods:** We used the UK Biobank and FinnGen, large datasets of health information and genotyping data. In UK Biobank (N = 502,131, N = 230,542 with linked GP records) we identified common infections in the linked health records for primary care and hospital admissions: skin infections (bacterial skin infections, fungal skin infections), respiratory infections (bacterial pneumonia, influenza, non-influenza viral respiratory tract infections), and urogenital infections (bacterial urinary tract infections, fungal genital infections). We assessed observational associations between BMI and infections using logistic regression. We used one-sample Mendelian randomisation to test for a causal effect of BMI on infections. We also used summary statistics from a BMI genome-wide association study, and infection outcomes from FinnGen (N = 500,348) to perform two-sample Mendelian randomisation and sensitivity analyses.

**Results:** 151,035 (65.5%) participants in UK Biobank with linked GP records had a record of infection in primary care, and 93,976 (18.7%) participants had a record of hospitalisation with infection.

Higher BMI was observationally associated with all infection types. One-sample MR demonstrates that higher BMI has a causal effect on skin infections in primary care (bacterial skin infections: Odds Ratio [OR] 1.37 [95%CI: 1.24-1.53] per 5 kg/m2 increase in BMI, p<0.001, fungal skin infections: OR 1.34 [95%CI: 1.18-1.53, p<0.001) and hospitalisation with skin infections (bacterial skin infections: OR 1.93 [95%CI: 1.71-2.19] per 5 kg/m^2^ increase in BMI, p<0.001, fungal skin infections: OR 2.81 [95%CI: 1.58-4.97, p<0.001). Two-sample MR provided further evidence of a causal effect of higher BMI on skin infections that is robust to pleiotropy. One-sample MR suggested evidence of a causal effect of higher BMI on some respiratory infections. However, two-sample MR sensitivity analyses suggest that this association is affected by pleiotropy. There was little evidence of a causal role of higher BMI in urogenital infections.

**Conclusions:** Mendelian randomisation provides strong evidence that higher BMI is a causal risk factor for bacterial and fungal skin infections. Weight loss interventions could help reduce the risk of mild and severe skin infections and be targeted to those at highest risk.

## Introduction

Infections are a major global health burden and remain a leading cause of mortality and morbidity. (1–3) Obesity is a global epidemic that affects over 1 in 8 people worldwide and continue to increase. (4–6) Higher BMI has been found to be associated with an increased risk of a range of common infections, including skin infections, respiratory infections, sepsis, and urinary tract infections. (7–10) This was further highlighted during the Covid-19 pandemic when increased BMI was found to be associated with poor infection outcomes such as hospitalisation. (11–13) However, risk factor associations reported for BMI have been almost entirely observational. There is limited evidence evaluating the causality of this association and so the potential causal role of BMI in infections is uncertain.

Understanding what causes infections is necessary to devise interventions to reduce risk. Despite infections being a major public health issue, there are currently few guidelines on how to reduce the risk of infections. BMI is a potentially modifiable risk factor and so, if found to be causal, provides an intervention target for reducing infection risk. This is particularly important in high-risk groups such as people with diabetes (14) and people with a history of recurrent or serious infection. (15)

Causal inference methods such as Mendelian randomisation (MR), can be used to test for causal relationships using observational data. MR is an epidemiological method that uses genetic variation as an unconfounded proxy for the exposure (a person’s genotype is randomly assigned at birth) to test for a causal relationship with an outcome. (16, 17) This method is becoming increasingly popular to overcome unmeasured confounding and reverse causality, major limitations in observational studies. (16–19)

There are few MR studies evaluating the causal role of modifiable risk factors for infections. One study assessing the association of BMI and infections has suggested a potential causal effect of BMI in hospitalisation for skin infections. (20) However, this study did not assess milder cases of infection treated in primary care, separate outcomes by infection aetiology, or validate results in two-sample MR. We therefore aimed to use an MR approach to test for a causal effect of higher BMI on common infections using large-scale population-based data.

## Methods

### Data

We used data from the UK Biobank and FinnGen, two of the largest datasets in the world combining genetic data and health information.

The UK Biobank is a large biomedical database containing health information and array-based genotyping data on around 500,000 participants. (21) The UK Biobank participants were volunteers recruited aged 40-70 between 2006 and 2010 across the UK. The baseline data are linked to electronic health record data for hospital admissions, and in a subset (n=230,542), primary care records.

FinnGen is a large-scale genomics initiative that has analysed over 500,000 Finnish biobank samples and correlated genetic variation with health data to understand disease mechanisms and predispositions. (22)

### Outcomes

We studied a range of infection outcomes covering the three body systems most commonly associated with hospitalisation with infections: skin infections (bacterial skin infections, fungal skin infections), respiratory infections (bacterial pneumonia, influenza, non-influenza viral respiratory tract infections), and urogenital infections (bacterial urinary tract infections, fungal genital infections). We also defined further subtypes of viral respiratory tract infections (lower respiratory tract infections, upper respiratory tract infections) and urinary tract infections (cystitis, pyelonephritis).

Using UK Biobank linked electronic health record datasets, for each of these infection types we defined two outcomes: infection treated in primary care, and hospitalisation with infection. This allowed us to evaluate the potential impact of BMI on both mild and severe infection outcomes. We defined primary care infections as any Read code for the infection type in the primary care dataset. We defined hospitalisation as an International Classification of Diseases 10^th^ Revision (ICD-10) code for the infection type as any diagnosis in the Hospital Episodes Statistics (HES) dataset. The Read and ICD-10 codes used to define each infection type are available in a Github repository: https://github.com/rhianhopkins/UKBiobank-MRInfections. Individuals without a diagnosis any of the studied infection types were used as controls in the respective analyses.

In FinnGen, we selected the closest matching outcomes to the ones we defined above in the UK Biobank. These were: L12_INFECT_SKIN (bacterial skin infections), AB1_DERMATOPHYTOSIS (fungal skin infection), J10_PNEUMOBACT (bacterial pneumonia), J10_INFLUENZA (influenza), J10_LOWERINF (lower respiratory tract infections), J10_UPPERINFEC (upper respiratory tract infections), N14_CYSTITIS (cystitis), and N14_PYELONEPHR (pyelonephritis). There was no closely matching outcome in FinnGen for fungal genital infections.

### Exposure

We defined BMI using the values recorded in the UK Biobank baseline assessment data. Individuals with missing values for BMI were excluded from the analyses (n=1,409).

The genetic variants used for BMI were obtained from a 2015 genome-wide association study (GWAS) of 339 224 individuals that reported 97 genome-wide significant loci. (23) We excluded sex-specific variants and those with potential pleiotropy or secondary signals within a locus, resulting in 72 variants used in our analysis. The study used to derive genetic instruments did not use the UK Biobank, which minimises the overlap of samples used for genotype-exposure and genotype-outcome associations. Supplementary table 1 lists the genetic variants used.

## Statistical methods

### Observational associations

We tested for observational associations between BMI and infection outcomes using a logistic regression model adjusted for age and sex in the UK Biobank cohort. Effect sizes were reported as odds ratios per 5 kg/m^2^ BMI increase.

### Mendelian randomisation assumptions

Mendelian randomisation methods rely on three core assumptions: (24)

1. Relevance: the genetic instrument is associated with the exposure
2. Exclusion restriction: the genetic instrument is not associated with confounders
3. Independence: the genetic instrument influences the outcome only through the exposure

Additionally, an important assumption is gene-environment equivalence (the genetic instrument influences an environmental exposure equivalently to a proposed intervention that changes the population distribution of the exposure). (25)

### One-sample Mendelian randomisation

We tested for a causal effect of BMI on infection using one-sample MR in the UK Biobank cohort. We first combined genetic variants for BMI into a genetic risk score (GRS) using the published effect size for each SNP as weights. We excluded individuals from the subsequent analyses if we could not calculate a genetic risk score. In the first stage of the two-stage least squares approach, the normalised GRS was regressed against BMI using a linear regression to derive genetically predicted exposure values. In the second stage, the genetically predicted exposure was regressed against the infection outcome in a logistic regression model, adjusted for age, sex, and the residuals. Effect sizes were reported as odds ratios on the same scale as the observational associations.

### Two-sample Mendelian randomisation

To further evaluate the causal role of BMI in infections, we also used two-sample Mendelian randomisation methods. This approach regressed the genotype-exposure effect sizes against the genotype-outcomes effect sizes for a set of SNPs associated with the exposure. We used inverse variance weighted (IVW) instrumental variable analysis and performed sensitivity analyses using additional methods that are more robust to potential violations of the standard MR assumptions (MR-Egger, median IV, penalised median IV). MR Egger is robust to pleiotropy and assesses whether genetic variants have pleiotropic effects on the outcome that differ on average from zero (indicated by the intercept). (26, 27) Median IV uses the median of the causal estimates for each genetic variant and allows up to 50% to be invalid instruments, and penalised median IV allows more precise causal estimates to contribute more weight to the analysis. (27, 28)

We obtained the effect sizes for the BMI genetic variants from the 2015 GWAS study described above (Supplementary table 1). To obtain genotype-outcome associations for these SNPs, we used publicly available summary statistics from FinnGen (22) for each of the infection outcomes.

Statistical analysis was performed using R version 4.3.0, and STROBE-MR reporting guidelines were followed. (29) R scripts used for analysis available at https://github.com/rhianhopkins/UKBiobank-MRInfections.

## Results

Of the 502,131 participants in the UK Biobank studied, 93,976 (18.7%) had a record of hospitalisation with infection. Of the 230,542 participants in the UK Biobank cohort who had linked GP records, 151,035 (65.5%) participants had a record of infection in primary care. The flow diagrams in Supplementary figure 1 and Supplementary figure 2 show the numbers of each infection outcome and controls for each analysis.

Table 1A and Table 1B describe the characteristics at UK Biobank assessment of those with and without infection, and numbers of participants with each infection type. Those with an infection in primary care were more commonly female and more commonly had diabetes than those without an infection. Those with a hospitalisation with infection were on average older, had slightly higher BMI, had greater deprivation, more commonly had diabetes and were more commonly smokers, compared to those without. Baseline characteristics by infection type are reported in Supplementary table 2A and Supplementary table 2B.

**Table 1A.** Baseline characteristics at UK Biobank assessment in individuals with linked GP data with and without a primary care infection.

|  | **Infection** | **No Infection** |
| --- | --- | --- |
| n | 151035 | 79507 |
| **Sex (%)** |  |  |
| Male | 64536 (42.7) | 39965 (50.3) |
| Female | 86499 (57.3) | 39542 (49.7) |
| **Age (mean (SD))** | 57.32 (8.02) | 56.45 (8.11) |
| **BMI (mean (SD))** | 27.75 (4.95) | 27.11 (4.54) |
| **Waist-hip-ratio (mean (SD))** | 0.87 (0.09) | 0.87 (0.09) |
| **Diagnosed diabetes (%)** |  |  |
| Yes | 8835 ( 5.8) | 3197 ( 4.0) |
| No | 141450 (93.7) | 75995 (95.6) |
| Unknown | 750 (0.5) | 315 (0.4) |
| **Townsend Deprivation Index (mean (SD))** | -1.34 (3.01) | -1.31 (3.08) |
| **Smoking status (%)** |  |  |
| Current | 16044 (10.6) | 8164 (10.3) |
| Previous | 52847 (35.0) | 26284 (33.1) |
| Never | 81288 (53.8) | 44701 (56.2) |
| Unknown | 856 ( 0.6) | 358 ( 0.5) |
| **Infection (%)** |  |  |
| Bacterial skin infection | 50271 (33.3) | NA |
| Fungal skin infection | 27748 (18.4) | NA |
| Bacterial pneumonia | 5716 ( 3.8) | NA |
| Influenza | 13594 ( 9.0) | NA |
| Non-influenza viral respiratory tract infection | 112984 (74.8) | NA |
| Lower respiratory tract infection | 76158 (50.4) | NA |
| Upper respiratory tract infection | 69680 (46.1) | NA |
| Bacterial urinary tract infection | 43115 (28.5) | NA |
| Cystitis | 42008 (27.8) | NA |
| Pyelonephritis | 2161 ( 1.4) | NA |
| Fungal genital infection | 17278 (11.4) | NA |

**Table 1B.** Baseline characteristics at UK Biobank assessment in individuals with and without an infection hospitalisation.

|  | **Infection** | **No Infection** |
| --- | --- | --- |
| n | 93976 | 408155 |
| **Sex (%)** |  |  |
| Male | 47002 (50.0) | 181972 (44.6) |
| Female | 46974 (50.0) | 226183 (55.4) |
| **Age (mean (SD))** | 59.67 (7.69) | 56.43 (8.06) |
| **BMI (mean (SD))** | 28.62 (5.50) | 27.16 (4.59) |
| **Waist-hip-ratio (mean (SD))** | 0.90 (0.09) | 0.87 (0.09) |
| **Diagnosed diabetes (%)** |  |  |
| Yes | 9784 (10.4) | 16602 ( 4.1) |
| No | 83494 (88.8) | 389636 (95.5) |
| Unknown | 698 (0.8) | 1917 (0.4) |
| **Townsend deprivation index (mean (SD))** | -0.82 (3.31) | -1.40 (3.03) |
| **Smoking Status (%)** |  |  |
| Current | 13607 (14.5) | 39330 ( 9.6) |
| Previous | 36554 (38.9) | 136366 (33.4) |
| Never | 42990 (45.7) | 230336 (56.4) |
| Unknown | 825 ( 0.9) | 2123 ( 0.5) |
| **Infection (%)** |  |  |
| Bacterial skin infection | 24433 (26.0) | NA |
| Fungal skin infection | 1059 ( 1.1) | NA |
| Bacterial pneumonia | 32074 (34.1) | NA |
| Influenza | 2005 ( 2.1) | NA |
| Non-influenza viral respiratory tract infection | 27346 (29.1) | NA |
| Lower respiratory tract infection | 22668 (24.1) | NA |
| Upper respiratory tract infection | 5612 ( 6.0) | NA |
| Bacterial urinary tract infection | 37900 (40.3) | NA |
| Cystitis | 6218 ( 6.6) | NA |
| Pyelonephritis | 3505 ( 3.7) | NA |
| Fungal genital infection | 796 ( 0.8) | NA |

### Mendelian randomisation demonstrates a causal effect of BMI on skin infections

Higher BMI was consistently observationally associated with skin infections, respiratory infections, and urogenital infections in primary care and hospitalisation with these infections (Figure 1A and Figure 1B).

**Figure 1.**
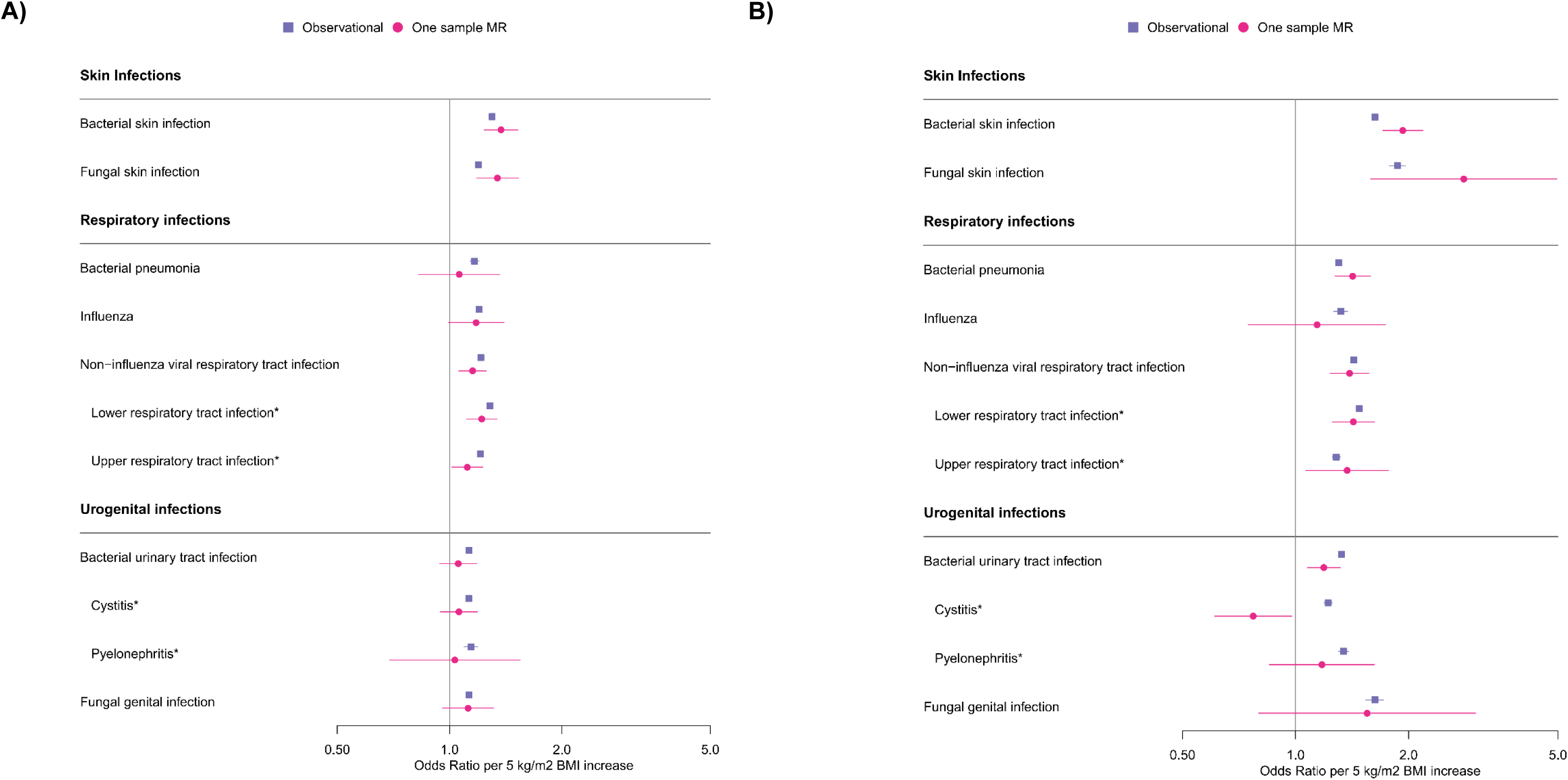
Forest plots of the observational (blue/purple squares) and one-sample Mendelian randomisation estimates (pink circles) for the association of BMI and A) infections in primary care and B) hospitalisation with infection. Odds ratios and 95% confidence intervals are given per 5 kg/m^2^. *Lower respiratory tract infection and upper respiratory tract infection are subgroups of non-influenza viral respiratory tract infection. Cystitis and pyelonephritis are subgroups of bacterial urinary tract infection.

One-sample Mendelian randomisation provides evidence that higher BMI has a causal effect on bacterial and fungal skin infections (Figure 1). A causal association with BMI was seen for skin infections both in primary care (bacterial skin infections: Odds Ratio [OR] 1.37 [95%CI: 1.24-1.53] per 5 kg/m^2^ increase in BMI, p<0.001, fungal skin infections: OR 1.34 [95%CI: 1.18-1.53, p<0.001, Figure 1B) and for hospitalisation with skin infections (bacterial skin infections: OR 1.93 [95%CI: 1.71-2.19] per 5 kg/m^2^ increase in BMI, p<0.001, fungal skin infections: OR 2.81 [95%CI: 1.58-4.97, p<0.001, Figure 1A). Two-sample Mendelian randomisation provides further evidence of a causal role of higher BMI in bacterial skin infections and fungal skin infections, and MR-Egger sensitivity suggests no evidence of pleiotropy (Figure 2A and Figure 2B).

**Figure 2.**
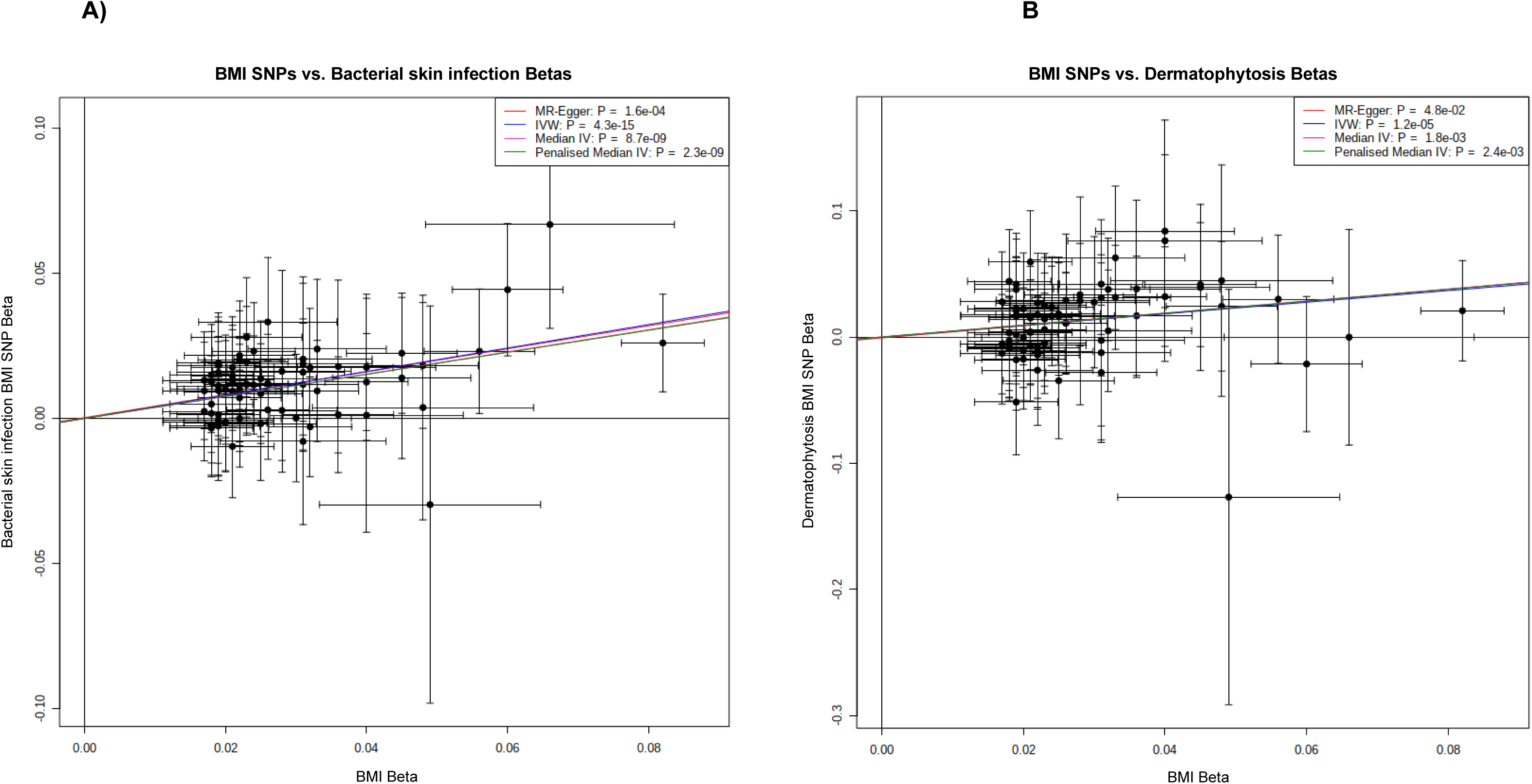
Two-sample Mendelian randomisation results for BMI and FinnGen skin infection outcomes. BMI SNP effect sizes shown against effect sizes for A) bacterial skin infections, and B) dermatophytosis (fungal skin infection). IVW, MR-Egger, median IV, and penalised median IV tests used.

One-sample Mendelian randomisation suggests a causal effect of BMI for some respiratory tract infections (Figure 1). Higher BMI was causally associated with non-influenza viral respiratory tract infections in primary care (OR 1.15 [95%CI 1.06-1.26], p=0.001) and for hospitalisation (OR 1.39 [95%CI: 1.24-1.57], p<0.001). Estimates for lower respiratory tract infections and upper respiratory tract infections were consistent with the overall association. There was also evidence of a causal association of higher BMI and hospitalisation with bacterial pneumonia (OR 1.42 [95%CI: 1.27-1.58], p<0.001), but not bacterial pneumonia in primary care. Two-sample Mendelian randomisation MR-Egger suggests evidence of horizontal pleiotropy in the association of higher BMI and respiratory infections (Supplementary figure 2).

One-sample Mendelian randomisation did not show consistent evidence of a causal effect of BMI on urogenital infections. An association was seen for hospitalisation with bacterial urinary tract infection (OR 1.19 [95%CI: 1.07-1.32], p <0.001); however, this was not consistent in primary care, or in the subgroup analyses of cystitis and pyelonephritis. Two-sample Mendelian randomisation also suggests little evidence of a causal effect of BMI on urogenital infections (Supplementary figure 3).

Full numeric results and numbers included in the analysis of each infection type are given in Supplementary table 3 for the observational analysis, and Supplementary table 4 for one-sample Mendelian randomisation. Supplementary table 5 provides full numerical results from two-sample Mendelian randomisation.

## Discussion

Using MR in large-scale population-based data, we demonstrate strong evidence of a causal role of increased BMI on bacterial and fungal infections. This causal effect was seen for milder cases of skin infection in primary care and hospitalisation with skin infections, and evidence was consistent in observational analyses and one- and two-sample MR. The effect sizes identified are highly clinically relevant, with a doubling of risk for hospitalisation with skin infection for every 5kg/m^2^ increase in BMI. Sensitivity analyses suggest this causal effect is robust to pleiotropy.

In observational and one-sample MR analyses we found evidence of a causal role of higher BMI on some respiratory infections. However, two-sample MR sensitivity analyses suggest that this association is affected by horizontal pleiotropy and the genetic variants may be influencing infections through a pathway outside of BMI. This therefore violates the core MR assumptions. We also found little evidence of a causal effect of BMI on urogenital infections.

### Interpretation/ Clinical implications

Given the significant morbidity and mortality due to infections, the evidence we have found of a causal effect of BMI on bacterial and fungal skin infections provides a potential important target for intervention. Our study suggests it is likely that weight loss can reduce the risk of these infections. As we found a causal association with infections in primary care and in hospital, this suggests that higher BMI could both cause increased risk of getting an infection and it being severe enough to be hospitalised. This evidence aligns with a previous study finding higher BMI was causally associated with increased risk of hospitalisation for skin infection. (20) Our study builds on this by providing evidence of a causal effect of higher BMI on milder cases of skin infection treated in primary care, stratifying by bacterial and fungal infection aetiologies, and validating using two-sample MR in a different population.

There are several plausible mechanisms which may explain why higher BMI causing infections. Intertrigo is a skin condition where increased skin folds result in areas of increased friction and moisture retention, making them susceptible to bacterial and fungal infections. (30, 31) Additionally, changes in blood flow within the skin due to obesity may impair the immune response to infections and lead to skin barrier impairment. (31) Skin conditions such as lymphoedema and venous insufficiency that are caused by obesity can also cause localised skin barrier breaches that can lead to infection. (30, 31)

The robust evidence we provide demonstrating a causal role of BMI in skin infections highlights that an emphasis on weight management is needed for skin infection prevention. While weight loss strategies need to be tailored to an individual, in the last few years there have been an increasing number of new medications that are available which can help weight loss, such as the next generation incretin-based agents. (32) Pharmacological intervention could therefore offer an effective intervention to support weight loss and improve infection outcomes. These weight loss interventions are expensive and therefore need to be targeted to those with the greatest benefit. Our findings suggest weight loss interventions could be particularly helpful to people who are most vulnerable to skin infections, for example people with diabetes (14) and people who have been admitted to hospital with a skin infection who are at high risk of being readmitted with another. (15)

### Strengths and limitations

A key strength of this study is the use of two of the largest datasets combining genetic data and health-related information that are available to study the causes of disease. The UK Biobank linked health record data allowed us to identify multiple common infection outcomes for a large cohort and the availability of systematically collected baseline data on BMI and genotyping data allowed us to test for causal associations using a one-sample MR approach. The existence of a large GWAS study for BMI provided valuable sources of genetic information for the study exposures. The availability of summary statistics for genotype associations with a range of infection types from FinnGen allowed us to further validate these associations using two-sample MR. The advantages of using MR to evaluate causal relationships is its robustness to confounding and reverse causation and can be used where conducting a randomised controlled trial is not possible. There are potential limitations to using data from the UK Biobank as the study recruited only individuals between the ages of 40-69 and there is a bias towards healthy individuals. The majority of UK Biobank are individuals of European ancestry and the numbers of individuals of other ancestries are too small to perform separate subgroup analyses in these groups. The summary statistics used in our two-sample MR also came from studies of Europeans only, and so associations may not be generalisable to other ancestries.

We robustly defined infection outcomes both using the primary care data and hospital records, which allowed us to test both if the risk factors were causing increased infections and if they were causing more severe infections requiring hospitalisation. We also carefully defined and separated infection outcomes by whether they were likely bacterial, viral, or fungal, which allowed us to test if the causal associations may differ by infection aetiology. In this study we comprehensively evaluated a large range of infection outcomes covering the most common types of infection leading to hospitalisation. We were unable to use two-sample MR to test for a causal association of BMI and fungal genital infections due to the lack of a close matching outcome in FinnGen, however one-sample MR suggested no evidence of a causal effect of higher BMI on this infection type. As with all studies involving health record data, misclassification of infection outcomes is possible and relies on correct coding in the records. In the UK hospital coding is performed by professional clinical coders and primary care coding usually performed by the clinician making the diagnosis. Using data from both sources mitigated against the impact of any systemic miscoding errors.

### Conclusion

MR demonstrates a causal role of increased BMI on bacterial and fungal skin infections. Interventions, such as the newly available medications for weight loss, could help lower the risk of skin infections. This is particularly important in people at greatly increased risk of infections, for example people with diabetes or a history of skin infection.

## Data Availability

All individual-level data used in this paper were obtained from the UK Biobank resource, and can be obtained from the UK Biobank at https://www.ukbiobank.ac.uk/enable-your-research/apply-for-access. Access to summary statistics from the FinnGen resource can be obtained at: https://www.finngen.fi/en/access_results/.

https://www.finngen.fi/en/access_results/

## Acknowledgements

This study was supported by the National Institute for Health and Care Research Exeter Biomedical Research Centre. The views expressed are those of the author(s) and not necessarily those of the NIHR or the Department of Health and Social Care. This study has been conducted using the UK Biobank Resource (project ID: 103356). We want to acknowledge the participants and investigators of the FinnGen study.

## Author contributions

The study concept and design were conceived and developed by RH, JMD, APM, and HDG. RH and EV undertook the analysis, with support from HDG. All authors provided support for the interpretation of results, critically revised the manuscript, and saw and approved the final article. HDG attests that all listed authors meet authorship criteria, that no others meeting the criteria have been omitted. HDG is responsible for the decision to submit for publication, and is the guarantor of this work and, as such, had full access to all the data in the study and takes responsibility for the integrity of the data and the accuracy of the data analysis.

## Ethics/data approval

Ethics approval for the UK Biobank study was obtained from the North West Centre for Research Ethics Committee (11/NW/0382). (21) Written informed consent was obtained from all participants.

## Rights Retention

For the purpose of open access, the author has applied a Creative Commons Attribution (CC BY) licence to any Author Accepted Manuscript version arising from this submission.

## Supplementary tables/ figures

**Supplementary figure 1A.**
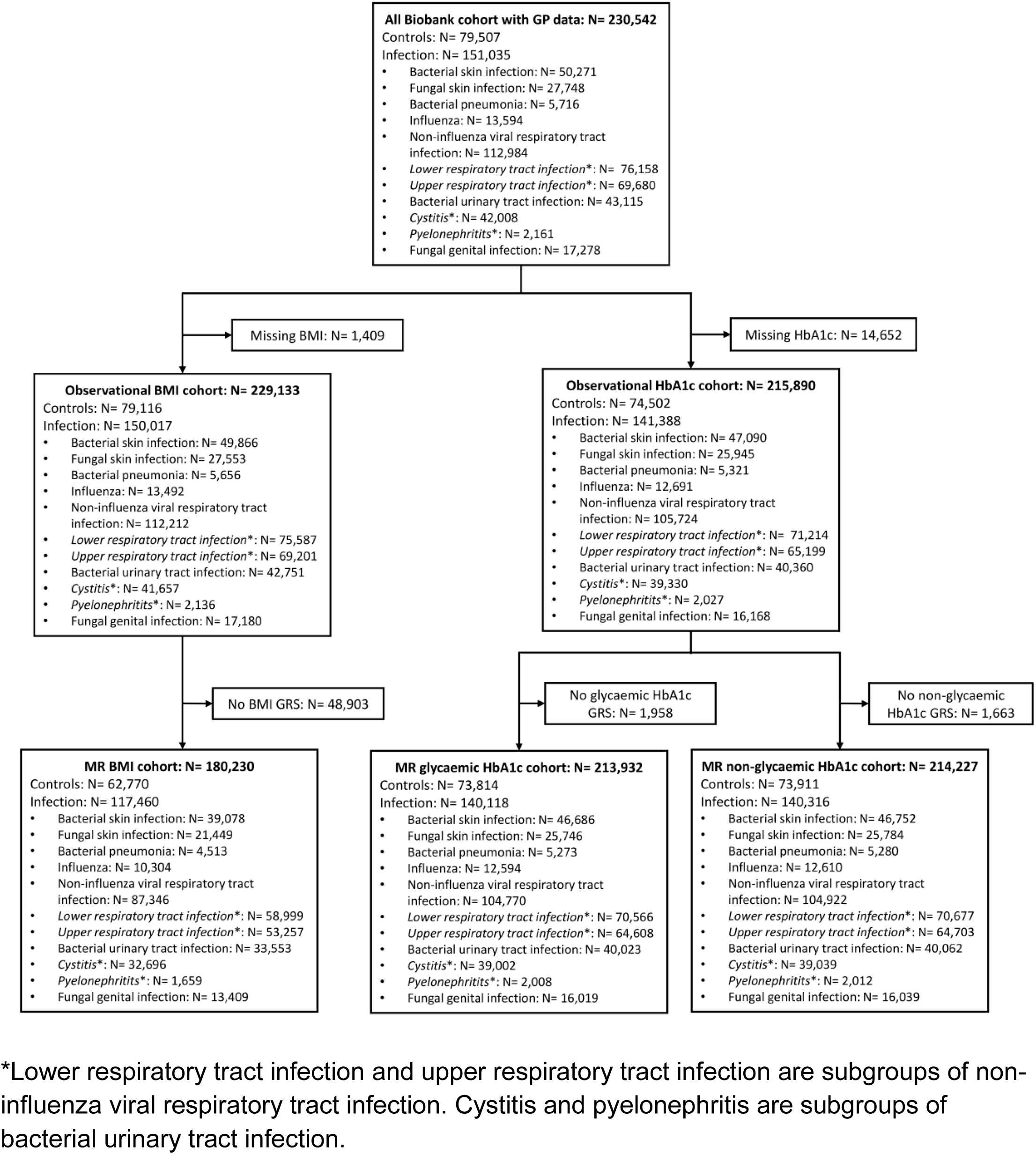
Study flow diagram of individuals with linked GP data from UK Biobank and numbers of primary care infection outcomes. *Lower respiratory tract infection and upper respiratory tract infection are subgroups of non-influenza viral respiratory tract infection. Cystitis and pyelonephritis are subgroups of bacterial urinary tract infection.

**Supplementary figure 1B.**
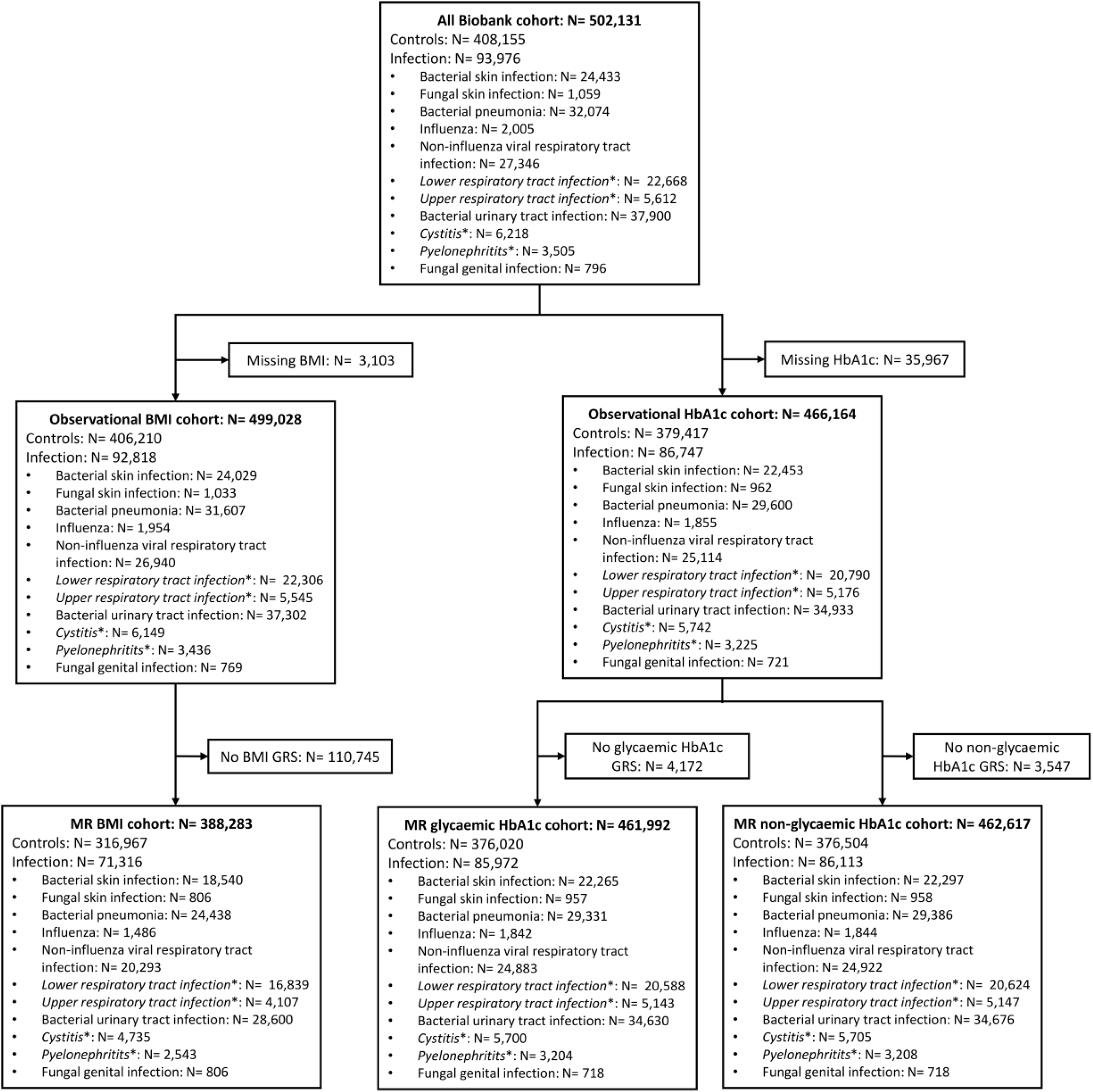
Study flow diagram of individuals from UK Biobank and numbers of hospitalisation with infection outcomes. *Lower respiratory tract infection and upper respiratory tract infection are subgroups of non-influenza viral respiratory tract infection. Cystitis and pyelonephritis are subgroups of bacterial urinary tract infection.

**Supplementary table 1.** Full list of BMI genetic variants used in Mendelian randomisation analysis.

| Trait | SNP | Trait raising | Other allele | BetaXG | seBetaXG |
| --- | --- | --- | --- | --- | --- |
| BMI | rs10733682 | A | G | 0.017 | 0.003 |
| BMI | rs11688816 | G | A | 0.017 | 0.003 |
| BMI | rs1808579 | C | T | 0.017 | 0.003 |
| BMI | rs6477694 | C | T | 0.017 | 0.003 |
| BMI | rs11583200 | C | T | 0.018 | 0.003 |
| BMI | rs12940622 | G | A | 0.018 | 0.003 |
| BMI | rs1528435 | T | C | 0.018 | 0.003 |
| BMI | rs29941 | G | A | 0.018 | 0.003 |
| BMI | rs3736485 | A | G | 0.018 | 0.003 |
| BMI | rs4740619 | T | C | 0.018 | 0.003 |
| BMI | rs1000940 | G | A | 0.019 | 0.003 |
| BMI | rs17724992 | A | G | 0.019 | 0.004 |
| BMI | rs1928295 | T | C | 0.019 | 0.003 |
| BMI | rs2033529 | G | A | 0.019 | 0.003 |
| BMI | rs2033732 | C | T | 0.019 | 0.004 |
| BMI | rs3849570 | A | C | 0.019 | 0.003 |
| BMI | rs6804842 | G | A | 0.019 | 0.003 |
| BMI | rs9400239 | C | T | 0.019 | 0.003 |
| BMI | rs9925964 | A | G | 0.019 | 0.003 |
| BMI | rs1167827 | G | A | 0.02 | 0.003 |
| BMI | rs2176598 | T | C | 0.02 | 0.004 |
| BMI | rs2365389 | C | T | 0.02 | 0.003 |
| BMI | rs2820292 | C | A | 0.02 | 0.003 |
| BMI | rs11126666 | A | G | 0.021 | 0.003 |
| BMI | rs12401738 | A | G | 0.021 | 0.003 |
| BMI | rs12885454 | C | A | 0.021 | 0.003 |
| BMI | rs2650492 | A | G | 0.021 | 0.004 |
| BMI | rs4256980 | G | C | 0.021 | 0.003 |
| BMI | rs11165643 | T | C | 0.022 | 0.003 |
| BMI | rs12286929 | G | A | 0.022 | 0.003 |
| BMI | rs17405819 | T | C | 0.022 | 0.003 |
| BMI | rs205262 | G | A | 0.022 | 0.004 |
| BMI | rs7243357 | T | G | 0.022 | 0.004 |
| BMI | rs7599312 | G | A | 0.022 | 0.003 |
| BMI | rs10132280 | C | A | 0.023 | 0.003 |
| BMI | rs1016287 | T | C | 0.023 | 0.003 |
| BMI | rs657452 | A | G | 0.023 | 0.003 |
| BMI | rs758747 | T | C | 0.023 | 0.004 |
| BMI | rs12566985 | G | A | 0.024 | 0.003 |
| BMI | rs7141420 | T | C | 0.024 | 0.003 |
| BMI | rs10968576 | G | A | 0.025 | 0.003 |
| BMI | rs17094222 | C | T | 0.025 | 0.004 |
| BMI | rs2121279 | T | C | 0.025 | 0.004 |
| BMI | rs2075650 | A | G | 0.026 | 0.005 |
| BMI | rs2112347 | T | G | 0.026 | 0.003 |
| BMI | rs3817334 | T | C | 0.026 | 0.003 |
| BMI | rs13191362 | A | G | 0.028 | 0.005 |
| BMI | rs3810291 | A | G | 0.028 | 0.004 |
| BMI | rs13078960 | G | T | 0.03 | 0.004 |
| BMI | rs10182181 | G | A | 0.031 | 0.003 |
| BMI | rs11057405 | G | A | 0.031 | 0.006 |
| BMI | rs11191560 | C | T | 0.031 | 0.005 |
| BMI | rs16951275 | T | C | 0.031 | 0.004 |
| BMI | rs17001654 | G | C | 0.031 | 0.005 |
| BMI | rs2245368 | C | T | 0.032 | 0.006 |
| BMI | rs7138803 | A | G | 0.032 | 0.003 |
| BMI | rs12429545 | A | G | 0.033 | 0.005 |
| BMI | rs3101336 | C | T | 0.033 | 0.003 |
| BMI | rs11727676 | T | C | 0.036 | 0.006 |
| BMI | rs2287019 | C | T | 0.036 | 0.004 |
| BMI | rs10938397 | G | A | 0.04 | 0.003 |
| BMI | rs12446632 | G | A | 0.04 | 0.005 |
| BMI | rs7899106 | G | A | 0.04 | 0.007 |
| BMI | rs1516725 | C | T | 0.045 | 0.005 |
| BMI | rs2207139 | G | A | 0.045 | 0.004 |
| BMI | rs16851483 | T | G | 0.048 | 0.008 |
| BMI | rs543874 | G | A | 0.048 | 0.004 |
| BMI | rs11847697 | T | C | 0.049 | 0.008 |
| BMI | rs6567160 | C | T | 0.056 | 0.004 |
| BMI | rs13021737 | G | A | 0.06 | 0.004 |
| BMI | rs17024393 | C | T | 0.066 | 0.009 |
| BMI | rs1558902 | A | T | 0.082 | 0.003 |

**Supplementary table 2A.** Baseline characteristics by infection type (primary care) in individuals with GP data in UK Biobank.

|  | No<br>Infectio<br>n | Bacterial<br>skin<br>infection | Fungal<br>skin<br>infection | Bacterial<br>pneumon<br>ia | Influenza | Other<br>viral<br>respirator<br>y<br>infection | Lower<br>respirator<br>y<br>infection | Upper<br>respirator<br>y<br>infection | Bacterial<br>urinary<br>tract<br>infection | Cystitis | Pyelonep<br>hritis | Fungal<br>genital<br>infection |
| --- | --- | --- | --- | --- | --- | --- | --- | --- | --- | --- | --- | --- |
| n | 79507 | 50271 | 27748 | 5716 | 13594 | 112984 | 76158 | 69680 | 43115 | 42008 | 2161 | 17278 |
| **Sex (%)** |  |  |  |  |  |  |  |  |  |  |  |  |
| Male | 39965<br>(50.3) | 23158<br>(46.1) | 15088<br>(54.4) | 2960<br>(51.8) | 5643<br>(41.5) | 47695<br>(42.2) | 32793<br>(43.1) | 26934<br>(38.7) | 9434<br>(21.9) | 9096<br>(21.7) | 430<br>(19.9) | 3732<br>(21.6) |
| Female | 39542<br>(49.7) | 27113<br>(53.9) | 12660<br>(45.6) | 2756<br>(48.2) | 7951<br>(58.5) | 65289<br>(57.8) | 43365<br>(56.9) | 42746<br>(61.3) | 33681<br>(78.1) | 32912<br>(78.3) | 1731<br>(80.1) | 13546<br>(78.4) |
| **Age (mean**<br>**(SD))** | 56.45<br>(8.11) | 57.46<br>(8.01) | 57.73<br>(7.89) | 58.93<br>(7.69) | 56.61<br>(7.92) | 57.42<br>(8.01) | 58.00<br>(7.89) | 56.95<br>(8.05) | 57.93<br>(7.91) | 57.94<br>(7.90) | 57.18<br>(8.06) | 55.57<br>(8.14) |
| **BMI (mean**<br>**(SD))** | 27.11<br>(4.54) | 28.37<br>(5.37) | 27.95<br>(4.89) | 27.85<br>(5.22) | 27.90<br>(5.01) | 27.94<br>(5.00) | 28.25<br>(5.13) | 27.90<br>(5.01) | 27.50<br>(5.10) | 27.49<br>(5.10) | 27.56<br>(5.32) | 27.43<br>(5.16) |
| **Waist-hip-**<br>**ratio (mean**<br>**(SD))** | 0.87<br>(0.09) | 0.88<br>(0.09) | 0.89<br>(0.09) | 0.89<br>(0.09) | 0.87<br>(0.09) | 0.87<br>(0.09) | 0.88<br>(0.09) | 0.87<br>(0.09) | 0.85<br>(0.09) | 0.85<br>(0.09) | 0.85<br>(0.09) | 0.84<br>(0.09) |
| **Diagnosed diabetes (%)** |  |  |  |  |  |  |  |  |  |  |  |  |
| Yes | 3197 (4.0) | 3747 (7.5) | 1808 (6.5) | 485 (8.5) | 809 (6.0) | 6887 (6.1) | 5240 (6.9) | 4089 (5.9) | 2593 (6.0) | 2495 (5.9) | 182 (8.4) | 1240 (7.2) |
| No | 75995<br>(95.6) | 46267<br>(92.0) | 25790<br>(92.9) | 5203<br>(91.0) | 12704<br>(93.5) | 105502<br>(93.4) | 70512<br>(92.6) | 65197<br>(93.6) | 40306<br>(93.5) | 39305<br>(93.6) | 1963<br>(90.8) | 15961<br>(92.4) |
| Unknown | 315<br>(0.4) | 257 (0.5) | 150 (0.6) | 28 (0.5) | 81 (0.5) | 595 (0.5) | 406 (0.5) | 394 (0.5) | 216 (0.5) | 208 (0.5) | 16 (0.8) | 77 (0.4) |
| **Townsend**<br>**deprivation**<br>**index (mean**<br>**(SD))** | -1.31<br>(3.08) | -1.29<br>(3.03) | -1.47<br>(2.96) | -1.14<br>(3.19) | -1.23<br>(3.06) | -1.31<br>(3.01) | -1.25<br>(3.04) | -1.32<br>(3.00) | -1.36<br>(2.99) | -1.37<br>(2.99) | -1.15<br>(3.08) | -1.41<br>(2.95) |
| **Smoking status (%)** |  |  |  |  |  |  |  |  |  |  |  |  |
| Current | 8164<br>(10.3) | 6127<br>(12.2) | 2687 (9.7) | 798<br>(14.0) | 1512<br>(11.1) | 12236<br>(10.8) | 9167<br>(12.0) | 6803 (9.8) | 4145 (9.6) | 4002 (9.5) | 254<br>(11.8) | 1629 (9.4) |
| Previous | 26284<br>(33.1) | 18046<br>(35.9) | 10083<br>(36.3) | 2223<br>(38.9) | 4651<br>(34.2) | 39913<br>(35.3) | 27753<br>(36.4) | 24036<br>(34.5) | 14828<br>(34.4) | 14407<br>(34.3) | 763<br>(35.3) | 5554<br>(32.1) |
| Never | 44701<br>(56.2) | 25808<br>(51.3) | 14800<br>(53.3) | 2660<br>(46.5) | 7343<br>(54.0) | 60151<br>(53.2) | 38770<br>(50.9) | 38408<br>(55.1) | 23880<br>(55.4) | 23342<br>(55.6) | 1128<br>(52.2) | 9993<br>(57.8) |
| Unknown | 358 ( 0.5) | 290 ( 0.6) | 178 ( 0.6) | 35 ( 0.6) | 88 ( 0.6) | 684 ( 0.6) | 468 ( 0.6) | 433 ( 0.6) | 262 ( 0.6) | 257 ( 0.6) | 16 ( 0.7) | 102 ( 0.6) |

**Supplementary table 2B.** Baseline characteristics by infection type (hospitalisation) in individuals in UK Biobank.

|  | No<br>Infectio<br>n | Bacterial<br>skin<br>infection | Fungal<br>skin<br>infection | Bacterial<br>pneumon<br>ia | Influenza | Other<br>viral<br>respirator<br>y<br>infection | Lower<br>respirator<br>y<br>infection | Upper<br>respirator<br>y<br>infection | Bacterial<br>urinary<br>tract<br>infection | Cystitis | Pyelonep<br>hritis | Fungal<br>genital<br>infection |
| --- | --- | --- | --- | --- | --- | --- | --- | --- | --- | --- | --- | --- |
| n | 408155 | 24433 | 1059 | 32074 | 2005 | 27346 | 22668 | 5612 | 37900 | 6218 | 3505 | 796 |
| **Sex (%)** |  |  |  |  |  |  |  |  |  |  |  |  |
| Male | 181972<br>(44.6) | 13366<br>(54.7) | 625<br>(59.0) | 18077<br>(56.4) | 961<br>(47.9) | 13874<br>(50.7) | 11933<br>(52.6) | 2355<br>(42.0) | 16607<br>(43.8) | 2473<br>(39.8) | 1215<br>(34.7) | 168<br>(21.1) |
| Female | 226183<br>(55.4) | 11067<br>(45.3) | 434<br>(41.0) | 13997<br>(43.6) | 1044<br>(52.1) | 13472<br>(49.3) | 10735<br>(47.4) | 3257<br>(58.0) | 21293<br>(56.2) | 3745<br>(60.2) | 2290<br>(65.3) | 628<br>(78.9) |
| **Age (mean**<br>**(SD))** | 56.43<br>(8.06) | 58.74<br>(7.91) | 59.82<br>(7.26) | 61.11<br>(7.06) | 59.68<br>(7.79) | 59.57<br>(7.83) | 60.26<br>(7.53) | 56.66<br>(8.35) | 60.53<br>(7.37) | 59.84<br>(7.60) | 58.05<br>(7.99) | 57.71<br>(8.85) |
| **BMI (mean**<br>**(SD))** | 27.16<br>(4.59) | 29.84<br>(6.24) | 30.72<br>(6.50) | 28.49<br>(5.56) | 28.51<br>(5.64) | 28.97<br>(5.65) | 29.15<br>(5.71) | 28.36<br>(5.45) | 28.55<br>(5.43) | 28.10<br>(5.10) | 28.60<br>(5.74) | 30.01<br>(6.71) |
| **Waist-hip-**<br>**ratio (mean**<br>**(SD))** | 0.87<br>(0.09) | 0.91<br>(0.09) | 0.92<br>(0.09) | 0.91<br>(0.09) | 0.90<br>(0.09) | 0.90<br>(0.09) | 0.91<br>(0.09) | 0.88<br>(0.09) | 0.89<br>(0.09) | 0.88<br>(0.10) | 0.88<br>(0.09) | 0.88<br>(0.09) |
| **Diagnosed diabetes (%)** |  |  |  |  |  |  |  |  |  |  |  |  |
| Yes | 16602<br>( 4.1) | 3287<br>(13.5) | 151<br>(14.3) | 3835<br>(12.0) | 244<br>(12.2) | 3220<br>(11.8) | 2884<br>(12.7) | 461 ( 8.2) | 4435<br>(11.7) | 547 ( 8.8) | 440<br>(12.6) | 154<br>(19.3) |
| No | 389636<br>(95.5) | 20955<br>(85.8) | 899<br>(84.9) | 27986<br>(87.3) | 1746<br>(87.1) | 23872<br>(87.3) | 19568<br>(86.3) | 5104<br>(90.9) | 33173<br>(87.5) | 5636<br>(90.6) | 3032<br>(86.5) | 632<br>(79.4) |
| Unknown | 1917<br>(0.4) | 191 (0.7) | 9 (0.8) | 253 (0.7) | 15 (0.7) | 254 (0.9) | 216 (1.0) | 47 (0.9) | 292 (0.8) | 35 (0.6) | 33 (0.9) | 10 (1.3) |
| **Townsend**<br>**deprivation**<br>**index (mean**<br>**(SD))** | -1.40<br>(3.03) | -0.67<br>(3.35) | -0.59<br>(3.30) | -0.63<br>(3.37) | -0.48<br>(3.46) | -0.58<br>(3.39) | -0.53<br>(3.40) | -0.72<br>(3.34) | -0.87<br>(3.30) | -1.00<br>(3.26) | -0.65<br>(3.33) | -0.53<br>(3.35) |
| **Smoking status (%)** |  |  |  |  |  |  |  |  |  |  |  |  |
| Current | 39330<br>( 9.6) | 3613<br>(14.8) | 155<br>(14.6) | 5934<br>(18.5) | 363<br>(18.1) | 4058<br>(14.8) | 3491<br>(15.4) | 699<br>(12.5) | 4644<br>(12.3) | 703<br>(11.3) | 463<br>(13.2) | 123<br>(15.5) |
| Previous | 136366<br>(33.4) | 9308<br>(38.1) | 387<br>(36.5) | 13303<br>(41.5) | 737<br>(36.8) | 10914<br>(39.9) | 9205<br>(40.6) | 2071<br>(36.9) | 14860<br>(39.2) | 2482<br>(39.9) | 1307<br>(37.3) | 295<br>(37.1) |
| Never | 230336<br>(56.4) | 11290<br>(46.2) | 504<br>(47.6) | 12533<br>(39.1) | 888<br>(44.3) | 12085<br>(44.2) | 9716<br>(42.9) | 2800<br>(49.9) | 18030<br>(47.6) | 2993<br>(48.1) | 1689<br>(48.2) | 367<br>(46.1) |
| Unknown<br>smoking | 2123 ( 0.5) | 222 ( 0.9) | 13 ( 1.2) | 304 ( 0.9) | 17 ( 0.8) | 289 ( 1.1) | 256 ( 1.1) | 42 ( 0.7) | 366 ( 1.0) | 40 ( 0.6) | 46 ( 1.3) | 11 ( 1.4) |

**Supplementary figure 2.**
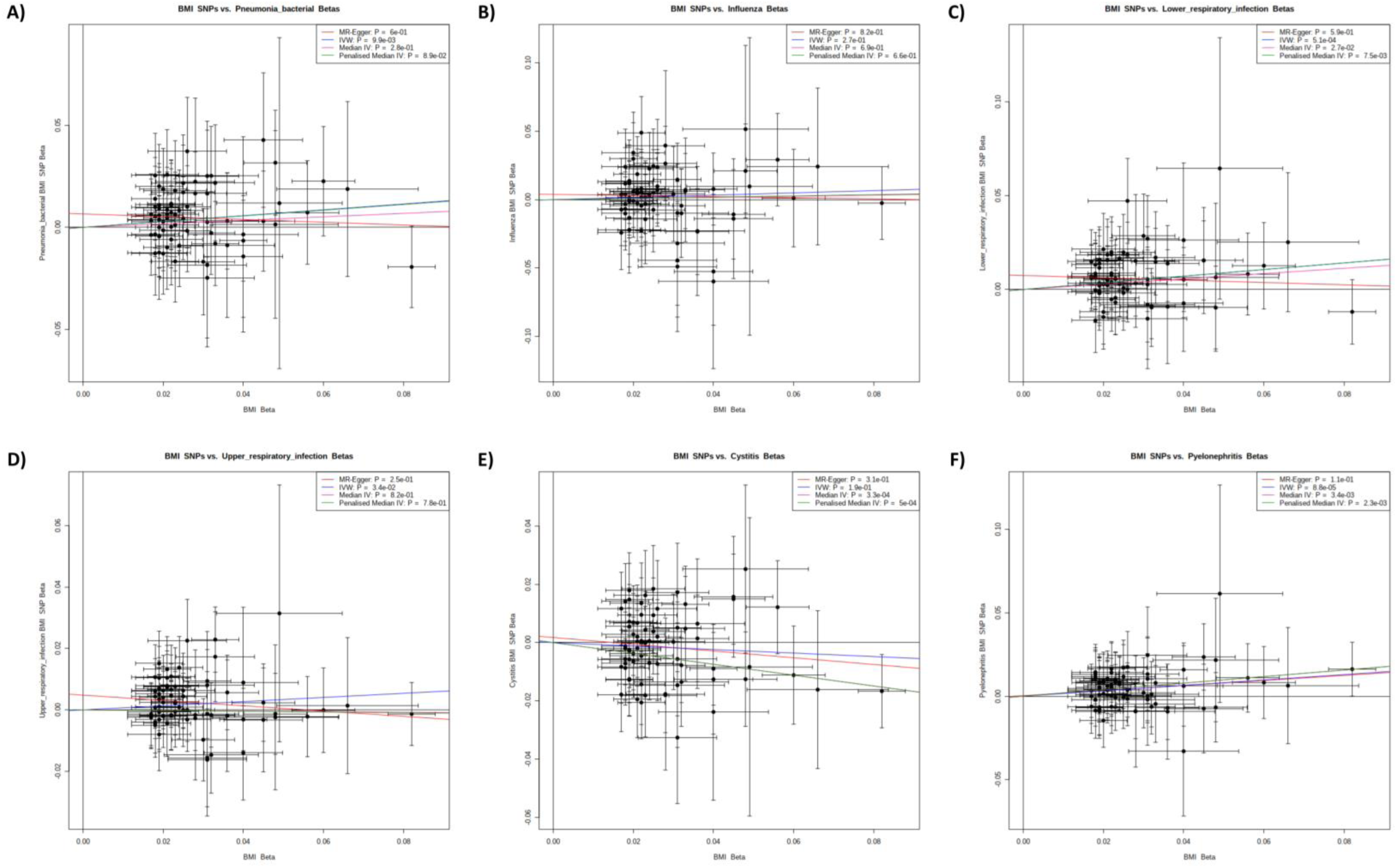
Two-sample Mendelian randomisation results for BMI and FinnGen infection outcomes. BMI SNP effect sizes shown against effect sizes for A) bacterial pneumonia, B) influenza, C) lower respiratory tract infections, D) upper respiratory tract infections, E) cystitis, and F) pyelonephritis. IVW, MR-Egger, median IV, and penalised median IV tests used.

**Supplementary table 3.** Observational associations of BMI and infections in primary and infection hospitalisation.

| Infection | Primary care |  |  | Hospitalisation |  |  |
| --- | --- | --- | --- | --- | --- | --- |
|  | N | Odds ratio (95%CI) per 5 kg/m<sup>2</sup> | p value | N | Odds ratio (95%CI) per 5 kg/m<sup>2</sup> | p value |
| Bacterial skin infection | 49866 | 1.30 (1.29-1.32) | <0.001 | 24029 | 1.63 (1.61-1.65) | <0.001 |
| Fungal skin infection | 27553 | 1.19 (1.18-1.21) | <0.001 | 1033 | 1.87 (1.78-1.96) | <0.001 |
| Bacterial pneumonia | 5656 | 1.17 (1.13-1.20) | <0.001 | 31607 | 1.30 (1.29-1.32) | <0.001 |
| Influenza | 13492 | 1.20 (1.18-2.22) | <0.001 | 1954 | 1.32 (1.26-1.38) | <0.001 |
| Non-influenza viral respiratory tract infection | 112212 | 1.21 (1.20-1.23) | <0.001 | 26940 | 1.43 (1.41-1.45) | <0.001 |
| Lower respiratory tract infection | 75587 | 1.28 (1.27-1.30) | <0.001 | 22306 | 1.48 (1.46-1.50) | <0.001 |
| Upper respiratory tract infection | 69201 | 1.21 (1.20-1.22) | <0.001 | 5545 | 1.28 (1.25-1.32) | <0.001 |
| Bacterial urinary tract infection | 42751 | 1.13 (1.11-1.14) | <0.001 | 37302 | 1.32 (1.31-1.34) | <0.001 |
| Cystitis | 41657 | 1.13 (1.11-1.14) | <0.001 | 6149 | 1.22 (1.19-1.25) | <0.001 |
| Pyelonephritis | 2136 | 1.14 (1.09-1.19) | <0.001 | 3436 | 1.34 (1.30-1.38) | <0.001 |
| Fungal genital infection | 17180 | 1.13 (1.11-1.15) | <0.001 | 769 | 1.63 (1.54-1.72) | <0.001 |

**Supplementary table 4.** One-sample Mendelian randomisation associations of BMI on infections in primary and infection hospitalisation.

| Infection | Primary care |  |  | Hospitalisation |  |  |
| --- | --- | --- | --- | --- | --- | --- |
|  | N | Odds ratio (95%CI) per 5 kg/m<sup>2</sup> | p value | N | Odds ratio (95%CI) per 5 kg/m<sup>2</sup> | p value |
| Bacterial skin infection | 39078 | 1.37 (1.24-1.53) | <0.001 | 18540 | 1.93 (1.71-2.19) | <0.001 |
| Fungal skin infection | 21449 | 1.34 (1.18-1.53) | <0.001 | 806 | 2.81 (1.58-4.97) | <0.001 |
| Bacterial pneumonia | 4513 | 1.06 (0.83-1.36) | 0.641 | 24438 | 1.42 (1.27-1.58) | <0.001 |
| Influenza | 10304 | 1.18 (0.99-1.40) | 0.062 | 1486 | 1.14 (0.75-1.74) | 0.538 |
| Non-influenza viral respiratory tract infection | 87346 | 1.15 (1.06-1.26) | 0.001 | 20293 | 1.39 (1.24-1.57) | <0.001 |
| Lower respiratory tract infection | 58999 | 1.22 (1.11-1.34) | <0.001 | 16839 | 1.43 (1.25-1.62) | <0.001 |
| Upper respiratory tract infection | 53257 | 1.12 (1.01-1.23) | 0.027 | 4107 | 1.37 (1.06-1.77) | 0.015 |
| Bacterial urinary tract infection | 33553 | 1.06 (0.94-1.18) | 0.359 | 28600 | 1.19 (1.07-1.32) | <0.001 |
| Cystitis | 32696 | 1.06 (0.94-1.19) | 0.329 | 4735 | 0.77 (0.61-0.98) | 0.032 |
| Pyelonephritis | 1659 | 1.03 (0.69-1.55) | 0.874 | 2543 | 1.18 (0.85-1.62) | 0.326 |
| Fungal genital infection | 13409 | 1.12 (0.96-1.31) | 0.156 | 592 | 1.55 (0.80-3.02) | 0.196 |

**Supplementary table 5.** Two-sample Mendelian randomisation results for BMI on infections. IVW, MR-Egger, median IV, and penalised median IV tests used.

| Trait | betal<br>VW2 | sebe<br>taIV<br>W2 | tIVW | pIV<br>W | IVR_<br>int | p.he<br>tero | beta<br>Egg<br>er | sebe<br>taEg<br>ger | tegg<br>er | pEg<br>ger | egge<br>r_int | int_<br>p | beta<br>WM | sebe<br>taW<br>M | tWM | pW<br>M | beta<br>PW<br>M | sebe<br>taP<br>WM | tPW<br>M | pPW<br>M | n_sn<br>p |
| --- | --- | --- | --- | --- | --- | --- | --- | --- | --- | --- | --- | --- | --- | --- | --- | --- | --- | --- | --- | --- | --- |
| Skin infection<br>(bacterial) | 0.40<br>4 | 0.04<br>1 | 9.98<br>0 | 0.00<br>0 | 0 | 0.72<br>2 | 0.39<br>6 | 0.09<br>9 | 3.99<br>1 | 0.00<br>0 | 0.00<br>0 | 0.91<br>8 | 0.38<br>2 | 0.06<br>6 | 5.75<br>4 | 0.00<br>0 | 0.38<br>0 | 0.06<br>4 | 5.97<br>2 | 0.00<br>0 | 71 |
| Dermatophytosis<br>(fungal skin<br>infection) | 0.46<br>0 | 0.09<br>8 | 4.71<br>3 | 0.00<br>0 | 0 | 0.39<br>4 | 0.48<br>3 | 0.24<br>0 | 2.00<br>9 | 0.04<br>8 | -<br>0.00<br>1 | 0.91<br>6 | 0.47<br>5 | 0.15<br>2 | 3.11<br>7 | 0.00<br>2 | 0.47<br>2 | 0.15<br>6 | 3.03<br>1 | 0.00<br>2 | 71 |
| Bacterial<br>pneumonia | 0.14<br>3 | 0.05<br>4 | 2.65<br>0 | 0.01<br>0 | 0 | 0.06<br>5 | -<br>0.06<br>8 | 0.13<br>0 | 0.52<br>2 | 0.60<br>4 | 0.00<br>7 | 0.08<br>0 | 0.08<br>6 | 0.08<br>0 | 1.07<br>1 | 0.28<br>4 | 0.14<br>0 | 0.08<br>2 | 1.69<br>8 | 0.08<br>9 | 71 |
| Influenza | 0.08<br>3 | 0.07<br>4 | 1.11<br>7 | 0.26<br>8 | 0 | 0.03<br>0 | -<br>0.04<br>2 | 0.18<br>2 | 0.23<br>0 | 0.81<br>9 | 0.00<br>4 | 0.45<br>5 | 0.04<br>1 | 0.10<br>3 | 0.40<br>1 | 0.68<br>8 | 0.04<br>6 | 0.10<br>6 | 0.43<br>6 | 0.66<br>3 | 71 |
| Lower<br>respiratory tract<br>infection | 0.17<br>7 | 0.04<br>9 | 3.64<br>3 | 0.00<br>1 | 0 | 0.01<br>8 | -<br>0.06<br>2 | 0.11<br>6 | 0.53<br>6 | 0.59<br>4 | 0.00<br>7 | 0.02<br>7 | 0.14<br>1 | 0.06<br>4 | 2.20<br>8 | 0.02<br>7 | 0.17<br>7 | 0.06<br>6 | 2.67<br>6 | 0.00<br>7 | 71 |
| Upper<br>respiratory tract<br>infection | 0.06<br>8 | 0.03<br>1 | 2.16<br>8 | 0.03<br>4 | 0 | 0.00<br>1 | -<br>0.08<br>6 | 0.07<br>4 | 1.16<br>4 | 0.24<br>8 | 0.00<br>5 | 0.02<br>6 | -<br>0.00<br>9 | 0.04<br>2 | 0.22<br>9 | 0.81<br>9 | -<br>0.01<br>2 | 0.04<br>1 | 0.28<br>3 | 0.77<br>7 | 71 |
| Cystitis | -<br>0.06<br>0 | 0.04<br>6 | 1.31<br>1 | 0.19<br>4 | 0 | 0.00<br>0 | -<br>0.11<br>6 | 0.11<br>2 | 1.02<br>9 | 0.30<br>7 | 0.00<br>2 | 0.58<br>8 | -<br>0.18<br>6 | 0.05<br>2 | 3.59<br>3 | 0.00<br>0 | -<br>0.18<br>6 | 0.05<br>4 | 3.48<br>2 | 0.00<br>0 | 71 |
| Pyelonephritis | 0.16<br>2 | 0.03<br>9 | 4.16<br>4 | 0.00<br>0 | 0 | 0.79<br>4 | 0.15<br>2 | 0.09<br>5 | 1.59<br>7 | 0.11<br>5 | 0.00<br>0 | 0.90<br>2 | 0.19<br>6 | 0.06<br>7 | 2.93<br>2 | 0.00<br>3 | 0.19<br>6 | 0.06<br>4 | 3.04<br>7 | 0.00<br>2 | 71 |

## Appendix

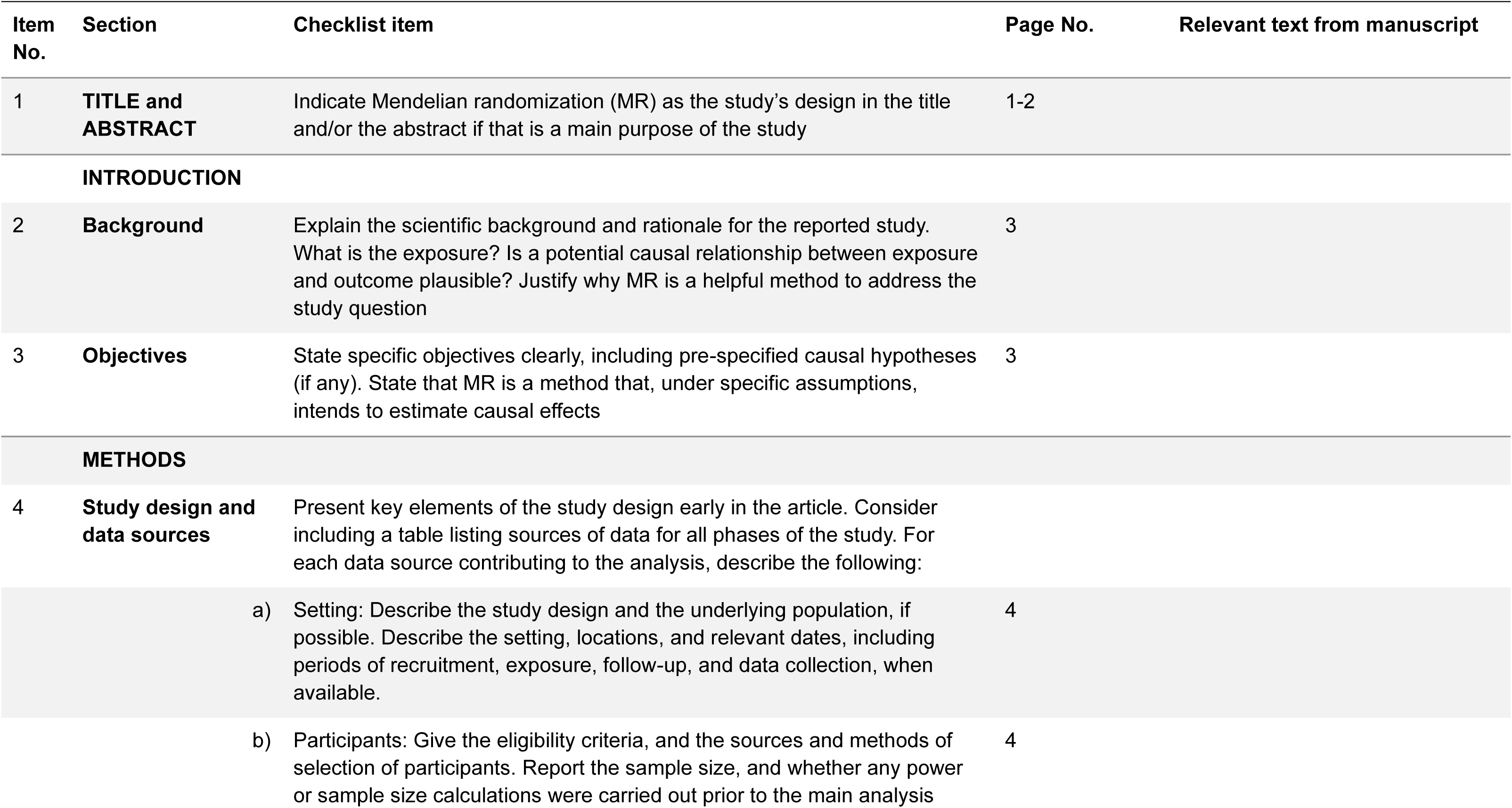
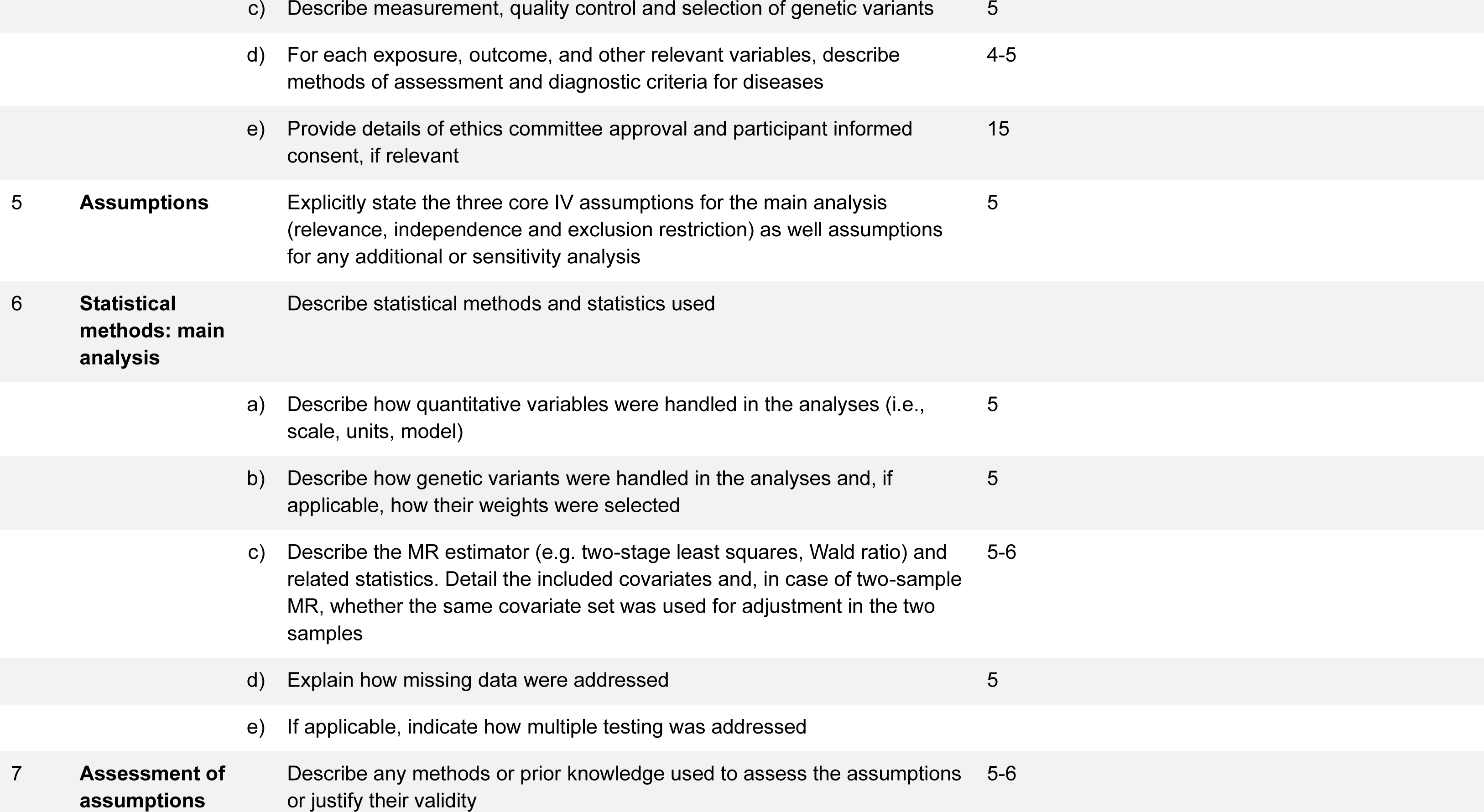
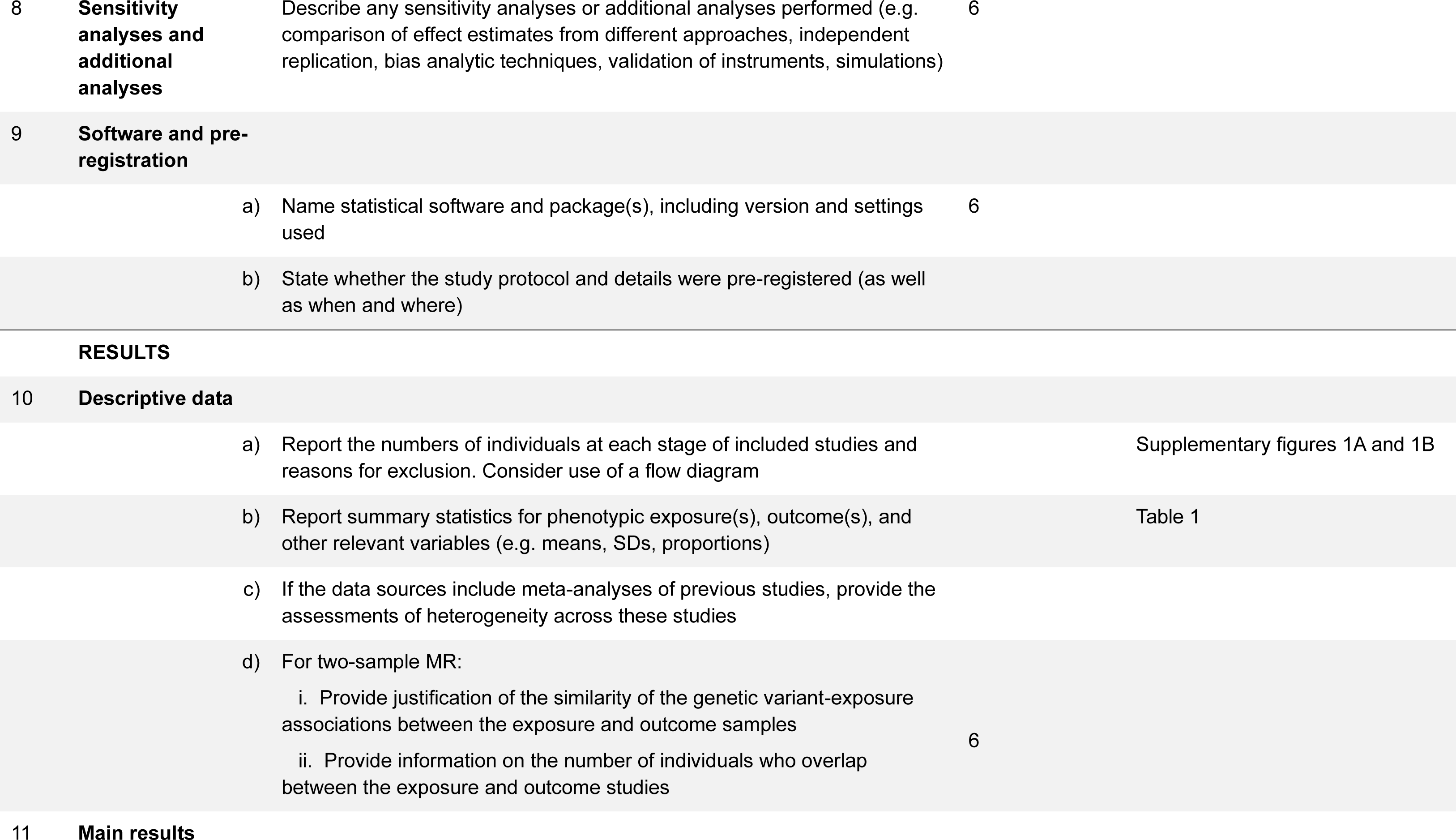
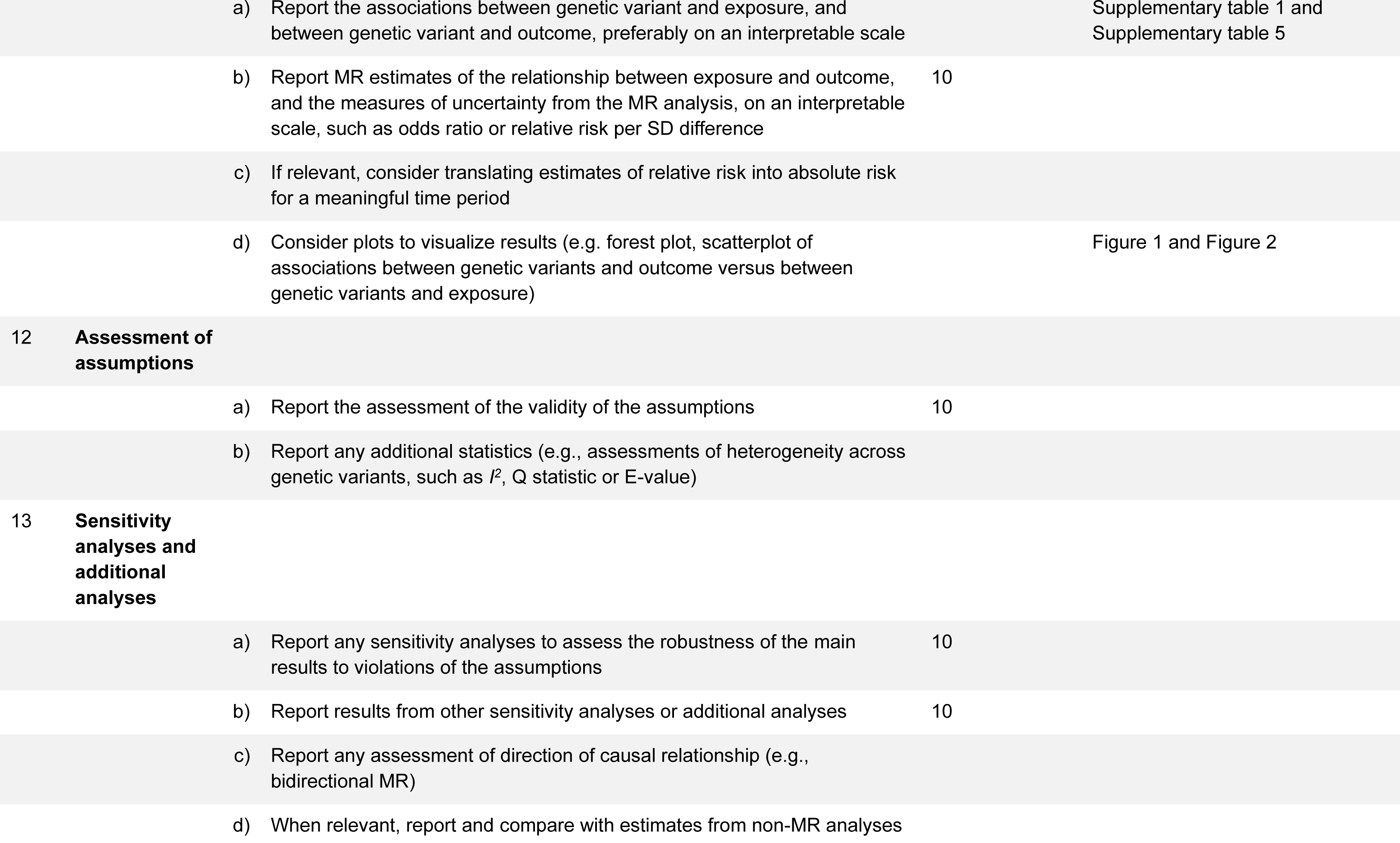
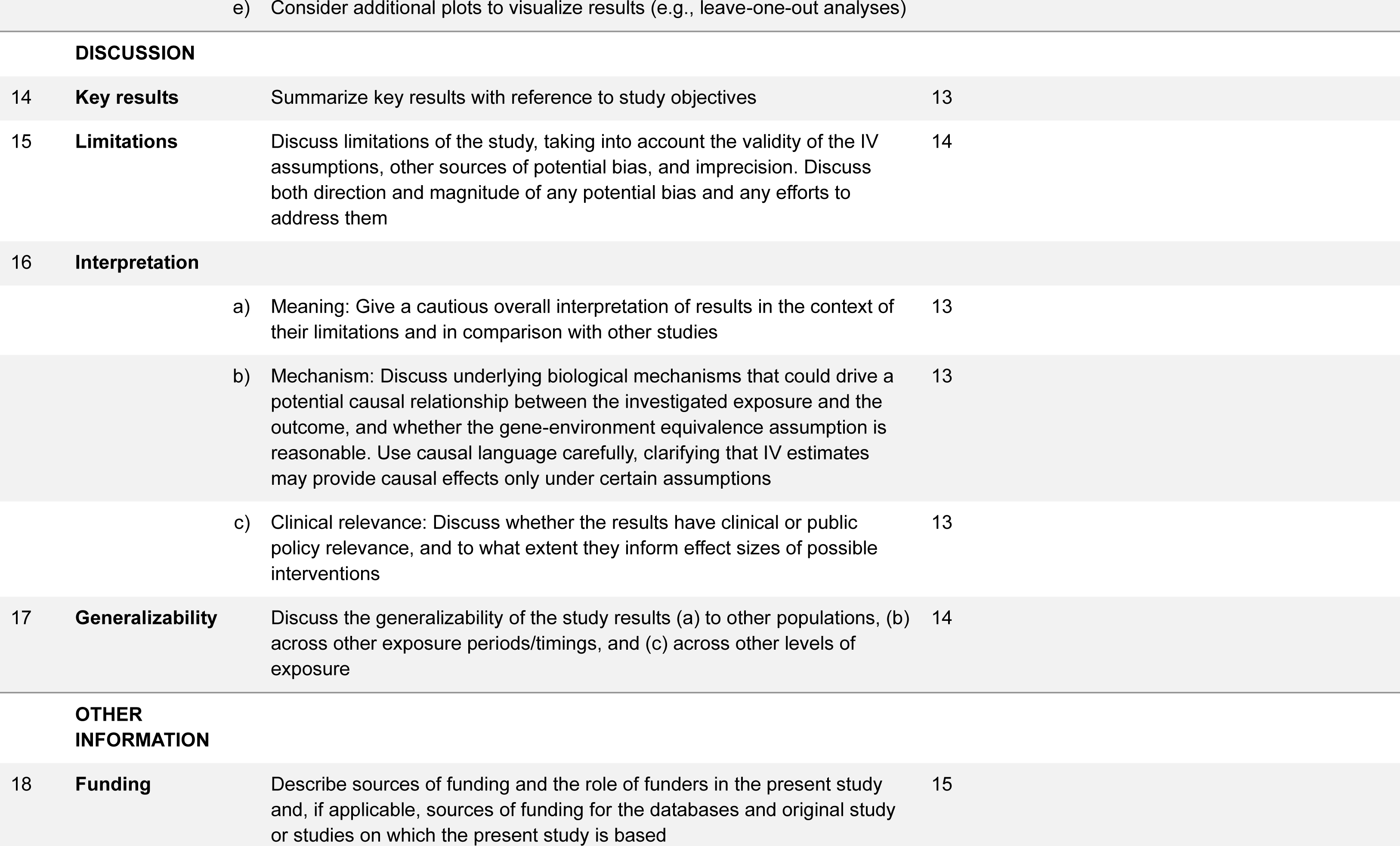
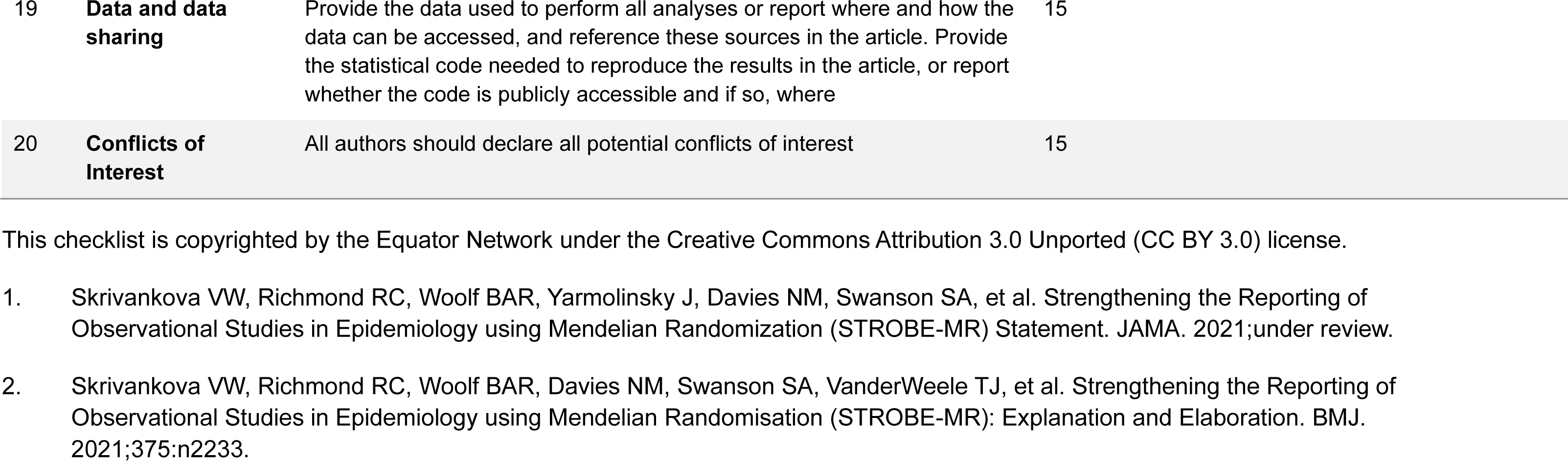
STROBE-MR checklist of recommended items to address in reports of Mendelian randomization studies^1 2^.

## References

1. Group IPC. Global burden associated with 85 pathogens in 2019: a systematic analysis for the Global Burden of Disease Study 2019. Lancet Infect Dis. 2024;24(8):868–95.

2. World Health Organisation. Global Health Estimates: Life expectancy and leading causes of death and disability 2021. Available from: https://www.who.int/data/gho/data/themes/mortality-and-global-health-estimates.

3. Collaborators GBDAR. Global mortality associated with 33 bacterial pathogens in 2019: a systematic analysis for the Global Burden of Disease Study 2019. Lancet. 2022;400(10369):2221–48.

4. Collaboration NCDRF. Worldwide trends in underweight and obesity from 1990 to 2022: a pooled analysis of 3663 population-representative studies with 222 million children, adolescents, and adults. Lancet. 2024;403(10431):1027–50.

5. World Health Organisation. Obesity and overweight 2024. Available from: https://www.who.int/news-room/fact-sheets/detail/obesity-and-overweight.

6. Collaborators GBDAB. Global, regional, and national prevalence of adult overweight and obesity, 1990-2021, with forecasts to 2050: a forecasting study for the Global Burden of Disease Study 2021. Lancet. 2025;405(10481):813–38.

7. Ghilotti F, Bellocco R, Ye W, Adami HO, Trolle Lagerros Y. Obesity and risk of infections: results from men and women in the Swedish National March Cohort. Int J Epidemiol. 2019;48(6):1783–94.

8. Harpsoe MC, Nielsen NM, Friis-Moller N, Andersson M, Wohlfahrt J, Linneberg A, et al. Body Mass Index and Risk of Infections Among Women in the Danish National Birth Cohort. Am J Epidemiol. 2016;183(11):1008–17.

9. Kaspersen KA, Pedersen OB, Petersen MS, Hjalgrim H, Rostgaard K, Moller BK, et al. Obesity and risk of infection: results from the Danish Blood Donor Study. Epidemiology. 2015;26(4):580–9.

10. Kornum JB, Norgaard M, Dethlefsen C, Due KM, Thomsen RW, Tjonneland A, et al. Obesity and risk of subsequent hospitalisation with pneumonia. Eur Respir J. 2010;36(6):1330–6.

11. Gao M, Piernas C, Astbury NM, Hippisley-Cox J, O’Rahilly S, Aveyard P, et al. Associations between body-mass index and COVID-19 severity in 6.9 million people in England: a prospective, community-based, cohort study. Lancet Diabetes Endocrinol. 2021;9(6):350–9.

12. Williamson EJ, Walker AJ, Bhaskaran K, Bacon S, Bates C, Morton CE, et al. Factors associated with COVID-19-related death using OpenSAFELY. Nature. 2020;584(7821):430–6.

13. Hopkins R, Young KG, Thomas NJ, Godwin J, Raja D, Mateen BA, et al. Risk factor associations for severe COVID-19, influenza and pneumonia in people with diabetes to inform future pandemic preparations: UK population-based cohort study. BMJ Open. 2024;14(1):e078135.

14. Carey IM, Critchley JA, DeWilde S, Harris T, Hosking FJ, Cook DG. Risk of Infection in Type 1 and Type 2 Diabetes Compared With the General Population: A Matched Cohort Study. Diabetes Care. 2018;41(3):513–21.

15. Karppelin M, Siljander T, Aittoniemi J, Hurme M, Huttunen R, Huhtala H, et al. Predictors of recurrent cellulitis in five years. Clinical risk factors and the role of PTX3 and CRP. J Infect. 2015;70(5):467–73.

16. Davey Smith G, Ebrahim S. What can mendelian randomisation tell us about modifiable behavioural and environmental exposures? BMJ. 2005;330(7499):1076-9.

17. Davies NM, Holmes MV, Davey Smith G. Reading Mendelian randomisation studies: a guide, glossary, and checklist for clinicians. BMJ. 2018;362:k601.

18. Stender S, Gellert-Kristensen H, Smith GD. Reclaiming mendelian randomization from the deluge of papers and misleading findings. Lipids Health Dis. 2024;23(1):286.

19. Burgess S, Butterworth A, Malarstig A, Thompson SG. Use of Mendelian randomisation to assess potential benefit of clinical intervention. BMJ. 2012;345:e7325.

20. Winter-Jensen M, Afzal S, Jess T, Nordestgaard BG, Allin KH. Body mass index and risk of infections: a Mendelian randomization study of 101,447 individuals. Eur J Epidemiol. 2020;35(4):347–54.

21. Bycroft C, Freeman C, Petkova D, Band G, Elliott LT, Sharp K, et al. The UK Biobank resource with deep phenotyping and genomic data. Nature. 2018;562(7726):203–9.

22. Kurki MI, Karjalainen J, Palta P, Sipila TP, Kristiansson K, Donner KM, et al. FinnGen provides genetic insights from a well-phenotyped isolated population. Nature. 2023;613(7944):508–18.

23. Locke AE, Kahali B, Berndt SI, Justice AE, Pers TH, Day FR, et al. Genetic studies of body mass index yield new insights for obesity biology. Nature. 2015;518(7538):197–206.

24. Boef AG, Dekkers OM, le Cessie S. Mendelian randomization studies: a review of the approaches used and the quality of reporting. Int J Epidemiol. 2015;44(2):496–511.

25. Burgess S, Woolf B, Mason AM, Ala-Korpela M, Gill D. Addressing the credibility crisis in Mendelian randomization. BMC Med. 2024;22(1):374.

26. Bowden J, Davey Smith G, Burgess S. Mendelian randomization with invalid instruments: effect estimation and bias detection through Egger regression. Int J Epidemiol. 2015;44(2):512–25.

27. Burgess S, Thompson SG. Interpreting findings from Mendelian randomization using the MR-Egger method. Eur J Epidemiol. 2017;32(5):377–89.

28. Bowden J, Davey Smith G, Haycock PC, Burgess S. Consistent Estimation in Mendelian Randomization with Some Invalid Instruments Using a Weighted Median Estimator. Genet Epidemiol. 2016;40(4):304–14.

29. Skrivankova VW, Richmond RC, Woolf BAR, Davies NM, Swanson SA, VanderWeele TJ, et al. Strengthening the reporting of observational studies in epidemiology using mendelian randomisation (STROBE-MR): explanation and elaboration. BMJ. 2021;375:n2233.

30. Pugliese G, Liccardi A, Graziadio C, Barrea L, Muscogiuri G, Colao A. Obesity and infectious diseases: pathophysiology and epidemiology of a double pandemic condition. Int J Obes (Lond). 2022;46(3):449–65.

31. Yosipovitch G, DeVore A, Dawn A. Obesity and the skin: skin physiology and skin manifestations of obesity. J Am Acad Dermatol. 2007;56(6):901–16; quiz 17-20.

32. Davies MJ, Aroda VR, Collins BS, Gabbay RA, Green J, Maruthur NM, et al. Management of Hyperglycemia in Type 2 Diabetes, 2022. A Consensus Report by the American Diabetes Association (ADA) and the European Association for the Study of Diabetes (EASD). Diabetes Care. 2022;45(11):2753–86.

